# Interim Safety and Immunogenicity of COVID-19 Omicron-BA.1 Variant-Containing Vaccine in Children

**DOI:** 10.1101/2023.06.23.23291767

**Authors:** Avika Dixit, Richard Bennett, Kashif Ali, Carl Griffin, Robert A. Clifford, Mark Turner, Rosanne Poston, Kelly Hautzinger, Anne Yeakey, Bethany Girard, Wen Zhou, Weiping Deng, Honghong Zhou, Sabine Schnyder Ghamloush, Barbara J. Kuter, Karen Slobod, Jacqueline M. Miller, Frances Priddy, Rituparna Das, the ROVER Study Investigators

## Abstract

**Objectives:** We report interim safety and immunogenicity results from a phase 3 study of omicron-BA.1 variant-containing (mRNA-1273.214) primary vaccination series (Part 1) and booster dose (Part 2) in children aged 6 months to 5 years (NCT05436834).

**Methods:** In Part 1, SARS-CoV-2 unvaccinated participants, including participants who received placebo in the KidCOVE study (NCT04796896), received 2 doses of mRNA-1273.214 (25-μg omicron-BA.1 and ancestral Wuhan-Hu-1 mRNA 1:1 co-formulation) primary series. In Part 2, participants who previously completed the mRNA-1273 (25-µg) primary series in KidCOVE received a mRNA-1273.214 (10-μg) booster dose. Primary objectives were safety, reactogenicity, and immunogenicity, including prespecified immune response success criteria.

**Results:** At the data cutoff (December 5, 2022), 179 participants had received ≥1 dose of mRNA-1273.214 primary series (Part 1) and 539 participants had received a mRNA-1273.214 booster dose (Part 2). The safety profile of mRNA-1273.214 primary series and booster dose was consistent with that of the mRNA-1273 primary series in this same age group, with no new safety concerns identified and no vaccine-related serious adverse events observed. Compared with neutralizing antibody responses induced by the mRNA-1273 primary series, both the mRNA-1273.214 primary series and booster elicited responses that were superior against omicron-BA.1 and non-inferior against ancestral Wuhan-Hu-1(D614G).

**Conclusions:** mRNA-1273.214 was immunogenic against BA.1 and D614G in children aged 6 months to 5 years, with a comparable safety profile to mRNA-1273, when given as a 2-dose primary series or as a booster dose after the mRNA-1273 primary series.

**Clinical Trial Registry:** NCT05436834

## Introduction

mRNA-based vaccines have been an essential approach to mitigate COVID-19 burden among children and adults worldwide.^1^ However, with the predominance of immune-evasive omicron sublineages globally,^2–4^ COVID-19 vaccines targeting the ancestral SARS-CoV-2 strain have shown reduced effectiveness.^5, 6^ A 2-dose mRNA-1273 primary series (SPIKEVAX; Moderna, Inc., Cambridge, USA) was well-tolerated in children aged 6 months to 11 years in the phase 2/3 KidCOVE trial, eliciting immune responses against the ancestral strain that were noninferior to those in young adults.^7, 8^ When the delta variant prevailed, the primary series demonstrated 88.0% (95% CI, 70.0-95.8; modified intent to treat [mITT]) efficacy in children aged 6-11 years.^8^ However, during omicron predominance, vaccine efficacy was 45.8% (95% CI, 19.0-63.4; mITT) and 39.6% (95% CI, 19.1-54.6; mITT) in children aged 6-23 months and 2-5 years, respectively.^7^ Consistent with clinical trial data, real-world effectiveness of 2 doses of mRNA-1273 among children aged 3-5 years is reported at 36% (95% CI, 15-52) during circulation of omicron sublineages.^9^ A higher infection and disease burden among children was observed after the emergence of omicron, with COVID-19 now the leading cause of infectious- and respiratory disease–related deaths among US individuals aged 0-19 years, surpassing influenza and pneumonia.^10^

Accordingly, COVID-19 vaccination strategies have been devised to broaden protection against SARS-CoV-2 variants. Variant-updated vaccines containing mRNAs encoding for both the ancestral SARS-CoV-2 strain (as in mRNA-1273) and omicron subvariant BA.1 (mRNA-1273.214) or BA.4/BA.5 (mRNA-1273.222) have been developed. In the United States, mRNA-1273.222 is authorized as a booster dose among individuals aged ≥6 months and as a primary series for age groups recommended to receive a primary series.^11^ In individuals aged ≥16 years, a booster dose of either variant-containing vaccine has elicited superior neutralizing antibody (nAb) responses against the relevant omicron strain with a safety profile similar to mRNA-1273.^12–14^ Given the consistent immunogenicity profile for the mRNA-1273 primary series between pediatric populations and young adults,^7, 8, 15^ variant-containing vaccination could provide similar benefits to children as seen in adults.

Here, we report interim safety and immunogenicity results from a phase 3 study of mRNA-1273.214 primary vaccination series (Part 1) and booster dose (Part 2) in children aged 6 months to 5 years, as a model for variant-containing COVID-19 vaccination in children.

## Methods

### Study Design and Participants

The open-label, phase 3 ROVER trial enrolled participants aged 6 months to 5 years at 24 US study sites and was conducted in 2 parts (NCT05436834; **Figure S1**). Part 1 enrolled participants who were not previously vaccinated against SARS-CoV-2, including participants who received placebo in KidCOVE (NCT04796896). Participants received 2 doses of mRNA-1273.214 (25-μg omicron-BA.1 and ancestral Wuhan-Hu-1 spike mRNA 1:1 co-formulation) primary series on Days 1 and 29, with a 12-month follow-up period after dose 2. Part 2 enrolled participants who previously completed mRNA-1273 (25-µg) primary series in KidCOVE. Participants received a mRNA-1273.214 (10-μg) booster ≥4 months after mRNA-1273 primary series and were followed for 6 months after booster. Participants who completed mRNA-1273 primary series in KidCOVE served as a historical comparator for inference of mRNA-1273.214 vaccine effectiveness.

Eligible participants were generally healthy or had stable chronic conditions, without a known SARS-CoV-2 infection in the preceding 90 days. Inclusion/exclusion criteria and study ethical statements are described the **Supplement**.

The protocol and study documents were approved before conduct of study procedures (Advarra Institutional Review Board). Written informed consent from parent(s)/legally authorized representative(s) of children was obtained before performing study procedures.

### Vaccines

mRNA-1273 contains mRNA encoding for the prefusion stabilized spike glycoprotein of the Wuhan-Hu-1 isolate of SARS-CoV-2. mRNA-1273.214 contains mRNAs (1:1 ratio) encoding for the prefusion stabilized spike glycoprotein of the Wuhan-Hu-1 isolate and for the omicron variant (B.1.1.529) subvariant BA.1 protein and was administered intramuscularly as a 25-µg primary series (Part 1; Days 1 and 29) or 10-µg booster (Part 2; booster Day 1).

### Objectives

The primary safety objective was to evaluate the safety and reactogenicity of mRNA-1273.214 administered as a 2-dose primary series (25-µg dose, Part 1) or as a single booster (10-µg dose after mRNA-1273 25-µg 2-dose primary series, Part 2) in participants aged 6 months to 5 years. Primary immunogenicity objectives were to infer effectiveness of mRNA-1273.214 based on immune responses against SARS-CoV-2 strains BA.1 and Wuhan-Hu-1 (D614G) measured 28 days after vaccination among participants (1) regardless of baseline SARS-CoV-2 status (Part 1) or (2) who completed mRNA-1273 primary series and were SARS-CoV-2 negative prior to booster (Part 2). Inference of mRNA-1273.214 effectiveness was based on assessment of immune responses in participants in the same age group after mRNA-1273 primary series in KidCOVE. Secondary objectives are described in the **Supplement**.

### Safety Assessments

Safety assessments included local and systemic solicited adverse reactions (SARs) through 7 days after vaccination; unsolicited adverse events (AEs) through 28 days after vaccination; and AEs leading to study withdrawal, medically attended AEs (MAAEs), AEs of special interest (AESIs; **Table S1**), and serious AEs (SAEs) from Day 1 through the interim analysis data cutoff.

### Immunogenicity Assessments

Blood samples for immunogenicity assessments were collected on Days 1 and 57 (Part 1) and booster Days 1 and 29 (Part 2). Samples on Days 1 and 57 after mRNA-1273 primary series from KidCOVE participants were evaluated as a historical comparator. nAb geometric mean concentrations (GMCs) against BA.1 and Wuhan-Hu-1 (D614G variant to optimize signal detection) were measured using pseudovirus neutralization assays.^16^ Seroresponse rates (SRRs) against BA.1 and D614G were evaluated.

### Statistical Analyses

Sample size calculations are presented in the **Supplement**. Safety analyses were based on the safety set, except for SARs which were based on the solicited safety set (**Table S2**). Part 1 primary immunogenicity analyses were assessed using the per-protocol immunogenicity set (PPIS); this analysis set was chosen as it was representative of the high prevalence of seropositivity among previously unvaccinated individuals. Part 2 primary immunogenicity analyses were conducted among participants in the PPIS with no serologic or virologic evidence of prior SARS-CoV-2 infection (PPIS-Neg).

nAb values with corresponding 95% CIs were calculated at each sampling timepoint. Number and percentage of participants with seroresponse after vaccination were provided with 2-sided 95% CIs (Clopper-Pearson method). For the primary immunogenicity analysis of nAb GMCs, a covariance model was utilized with the nAb value as the dependent variable and group (mRNA-1273.214 vs mRNA-1273) as the fixed variable, adjusted for age subgroup (6-23 months and 2-5 years). The Part 1 model also adjusted for baseline SARS-CoV-2 infection status (ie, evidence of prior SARS-CoV-2 infection). The nAb GMC value was estimated using the model geometric least square mean (GLSM). The geometric mean ratio (GMR; mRNA-1273.214 vs mRNA-1273) was estimated using the ratio of GLSM, with 2-sided 95% CI from the model employed to assess differences in immune response between the 2 groups. Superiority and non-inferiority were declared if the lower bound (LB) of the GMR 95% CI was >1 and >0.667, respectively. SRR analyses are described in the **Supplement**.

## Results

### Part 1

#### Participants

From enrollment to the interim analysis (June 21, 2022, to December 5, 2022), 179 participants received ≥1 dose of mRNA-1273.214 and 142 (79.3%) reached day 28 and received dose 2 (**Figure 1**). Median (interquartile range [IQR]) age of participants was 3 (1-3) years and most participants were White (65.4%; **Table 1**). Median (IQR) follow-up time was 85 (43-113) days after dose 1 and 68 (34-90) days after dose 2. Baseline SARS-CoV-2 positivity rate was 63.1%.

**Figure 1.**
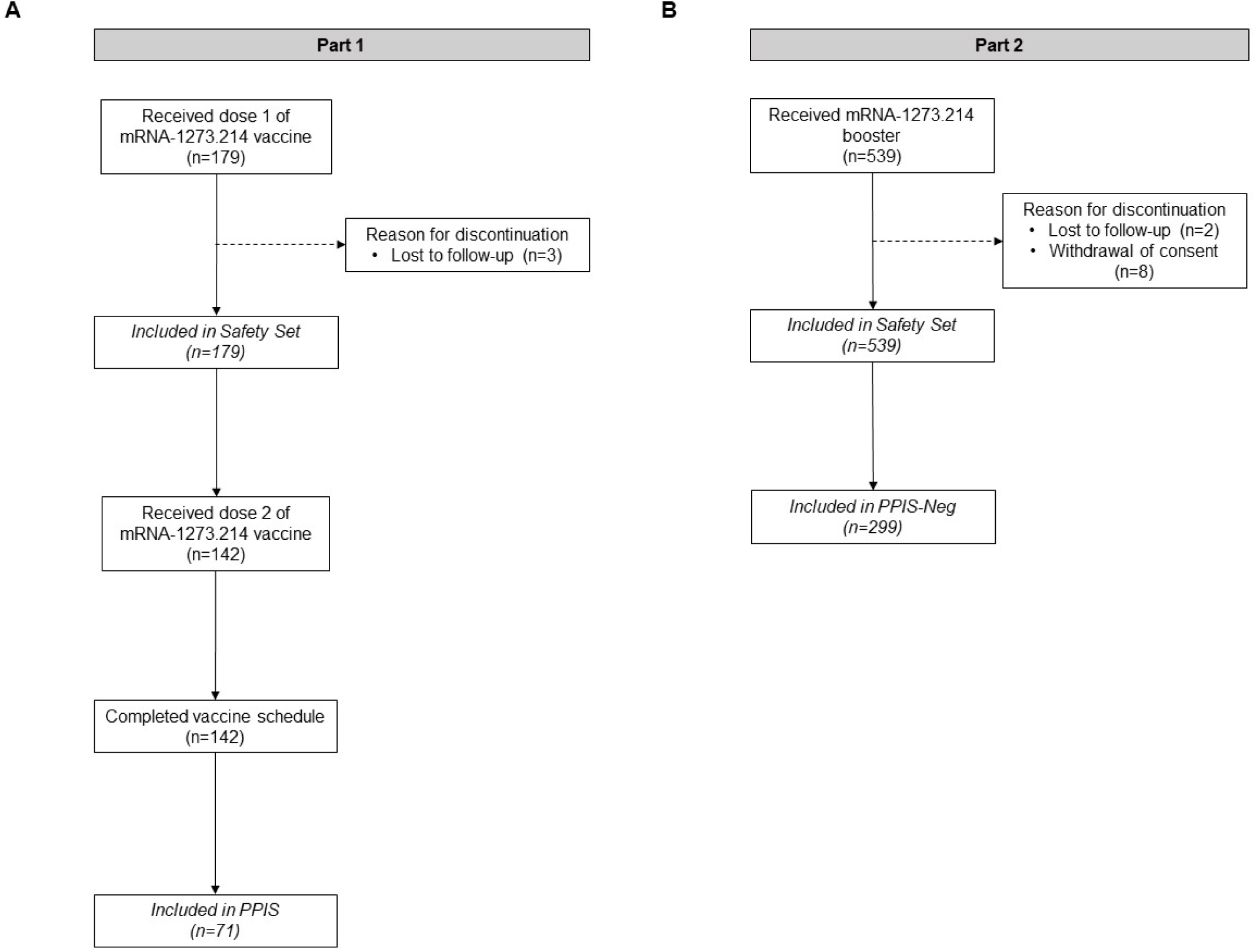
Participant disposition in Part 1 (A) and Part 2 (B). PPIS, per-protocol immunogenicity set; PPIS-Neg, per-protocol immunogenicity set-negative.

**Table 1.** Participant Demographics in Part 1 **(mRNA-1273.214 Primary Series; Safety Set)**

|  | **Part 1 Participants**<br>**(N=179)** |
| --- | --- |
| Mean age, years (SD) | 2.6 (1.4) |
| Median age, years (IQR; min-max) | 3.0 (1.0-3.0; 0.5-5.0) |
| Male, n (%) | 98 (54.7) |
| Race, n (%) |  |
| White | 117 (65.4) |
| Black | 46 (25.7) |
| Asian | 5 (2.8) |
| American Indian or Alaska Native | 1 (0.6) |
| Native Hawaiian or Other Pacific Islander | 0 |
| Multiracial | 8 (4.5) |
| Other | 2 (1.1) |
| Unknown or not reported | 0 |
| Ethnicity, n (%) |  |
| Hispanic or Latino | 21 (11.7) |
| Not Hispanic or Latino | 158 (88.3) |
| Unknown or not reported | 0 |
| Baseline SARS-CoV-2 infection, n (%)<sup>a</sup> |  |
| Positive | 113 (63.1) |
| Negative | 42 (23.5) |
| Missing | 24 (13.4) |

IQR, interquartile range; RT-PCR, reverse transcription-polymerase chain reaction.
<sup>a</sup>Baseline SARS-CoV-2 status was categorized as positive if there was immunologic or virologic evidence of prior COVID-19, defined as positive RT-PCR test or positive Elecsys result on or before Day 1. Negative was defined as negative RT-PCR test and negative Elecsys result on or before Day 1.

#### Safety

At least 1 SAR within 7 days after dose 1 and 2 was reported by 102 (57.0%) and 89 participants (63.1%), respectively (**Figure S2**). Most SARs were grade 1 or 2; no grade 4 events were reported. The most commonly reported local SAR after either injection was pain (51.4%). Irritability/crying (55.2%), sleepiness (43.7%), and fatigue (41.8%) were the most commonly reported systemic SARs. Fever (≥38⁰C/100.4⁰F) was reported by 16 participants (8.9%) after dose 1 and 19 (13.5%) after dose 2. Grade 3 fever was reported by 2 participants (1.1%) after dose 1 and 2 (1.4%) after dose 2. Median (range) time of SAR onset was 1.0 (1-5) and 1.0 (1-6) day after doses 1 and 2, respectively, and duration was 2.0 (1-7) days after doses 1 and 2.

Unsolicited AEs were reported by 30.7% of participants within 28 days after either injection; 1.1% were considered vaccine-related (1 report each of diarrhea and croup; **Table S3**). The most common unsolicited AEs were upper respiratory tract infection (8.9%), rhinorrhea (2.8%), and ear infection (2.2%). One participant (0.6%) experienced an SAE (asthma exacerbation 14 days after dose 1) considered unrelated to vaccination. There were no AESIs or AEs leading to discontinuation of study vaccine or study participation up to the data cutoff.

#### Immunogenicity

The primary series induced nAb levels against BA.1 that were superior to those elicited by mRNA-1273 in KidCOVE (**Figure 2A**). Among the PPIS, observed nAb GMCs (95% CI) against BA.1 were 1889.7 (1430.0-2497.2) after mRNA-1273.214 primary series and 74.3 (67.7-81.7) after mRNA-1273. The Day 57 estimated GMR (95% CI) was 25.4 (20.1-32.1), meeting the pre-specified superiority criterion. Among the PPIS-Neg, mRNA-1273.214 primary series induced higher observed nAb GMCs (95% CI) against BA.1 than mRNA-1273 primary series at Day 57 (mRNA-1273.214: 1037.9 [786.5-1369.7]; mRNA-1273: 65.7 [60.6-71.3]), with an estimated GMR (95% CI) of 15.8 (11.4-22.0). Day 57 SRRs (95% CI) for BA.1 were 89.3% (78.1-96.0) and 86.8% (82.0-90.7) after mRNA-1273.214 and mRNA-1273 primary series, respectively (**Table S4**).

**Figure 2.**
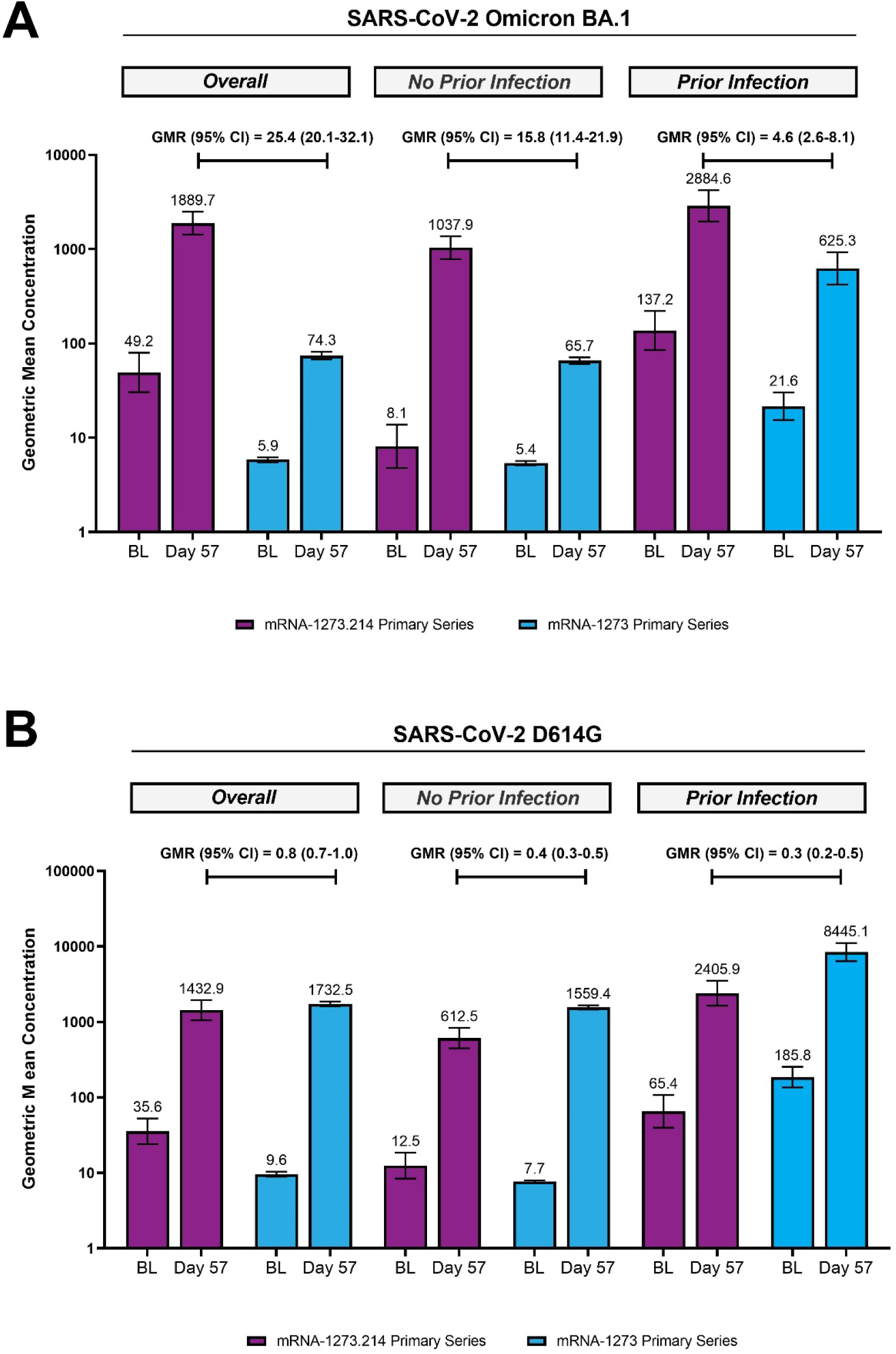
Serum neutralizing antibodies against BA.1 (A) and D614G (B) after mRNA-1273.214 primary series or mRNA-1273 primary series in Part 1. Pseudovirus nAbs at BL and at 28 days after dose 2 (day 57) are shown for participants in the PPIS and by baseline SARS-CoV-2 status. No prior SARS-CoV-2 infection was defined as negative RT-PCR test and negative Elecsys result on or before Day 1. Prior SARS-CoV-2 infection was indicated by a positive RT-PCR test or positive Elecsys result on or before Day 1. Overall: mRNA-1273.214, n=71; mRNA-1273, n=632; no prior infection: mRNA-1273.214, n=26; mRNA-1273, n=590; prior infection: mRNA-1273.214, n=45; mRNA-1273, n=42. Antibody values below the LLOQ were replaced by 0.5 x LLOQ. Values greater than the ULOQ were replaced by the ULOQ if actual values were not available. LLOQs were 8 (BA.1) or 10 (D614G); ULOQs were 41984 (BA.1) or 4505600 (D614G). BL, baseline; CI, confidence interval; GMC, geometric mean concentration; GMR, geometric mean ratio; LLOQ, lower limit of quantification; nAb, neutralizing antibody; PPIS, per-protocol immunogenicity set; RT-PCR, reverse transcription-polymerase chain reaction; ULOQ, upper limit of quantification.

At Day 57, the primary series elicited nAb responses against D614G that were non-inferior to those induced by mRNA-1273 primary series in KidCOVE (**Figure 2B**). Among the PPIS, the observed nAb GMCs (95% CI) against D614G were 1432.9 (1054.5-1947.0) after mRNA-1273.214 and 1732.5 (1611.5-1862.5) after mRNA-1273. The estimated GMR was 0.8 (95% CI, 0.7-1.0), meeting the pre-specified criterion of non-inferiority. Among the PPIS-Neg, the observed nAb GMCs (95% CI) against D614G were 612.5 (448.2-836.9) after mRNA-1273.214 and 1559.4 (1459.2-1666.6) after mRNA-1273, with an estimated GMR of 0.4 (95% CI, 0.3-0.5). Day 57 SRRs (95% CI) were 97.0% (89.5-99.6) and 99.5% (98.5-99.9) after mRNA-1273.214 and mRNA-1273 primary series, respectively (**Table S4**).

### Part 2

#### Participants

From enrollment to the interim analysis (June 22, 2022, to December 5, 2022), 539 participants received a mRNA-1273.214 booster (**Figure 1**). Median (IQR) age was 3 (2-4; the older age range reflecting the time lapse between primary series receipt in KidCOVE and become eligible for booster dosing) years and most participants were White (81.1%; **Table 2**). Median (IQR) follow-up after the mRNA-1273.214 booster was 117 (109-130) days and the median time between dose 2 of mRNA-1273 primary series and mRNA-1273.214 booster was 7.9 (7.0-8.3) months.

**Table 2.** Participant Demographics in Part 2 (**mRNA-1273.214 Booster; Safety Set**)

|  | **Part 2 Participants**<br>**(N=539)** |
| --- | --- |
| Mean age, years (SD) | 2.7 (1.3) |
| Median age, years (IQR; min-max) | 3.0 (2.0-4.0; 0.9-5.0) |
| Male (%) | 276 (51.2) |
| Race, n (%) |  |
| White | 437 (81.1) |
| Black | 17 (3.2) |
| Asian | 26 (4.8) |
| American Indian or Alaska Native | 0 |
| Native Hawaiian or Other Pacific Islander | 2 (0.4) |
| Multiracial | 52 (9.6) |
| Other | 0 |
| Unknown | 1 (0.2) |
| Not reported | 4 (0.7) |
| Ethnicity, n (%) |  |
| Hispanic or Latino | 59 (10.9) |
| Not Hispanic or Latino | 476 (88.3) |
| Unknown | 2 (0.4) |
| Not reported | 2 (0.4) |
| Pre-booster SARS-CoV-2 infection, n (%)<sup>a</sup> |  |
| Positive | 153 (28.4) |
| Negative | 331 (61.4) |
| Missing | 55 (10.2) |

IQR, interquartile range; RT-PCR, reverse transcription-polymerase chain reaction.
<sup>a</sup>Pre-booster SARS-CoV-2 status was categorized as positive if there was immunologic or
virologic evidence of prior COVID-19, defined as positive RT-PCR test or positive Elecsys
result on or before booster dose Day 1. Negative was defined as negative RT-PCR
test and negative Elecsys result on or before booster dose Day 1.

#### Safety

Overall, 382 participants (70.9%) reported ≥1 SAR within 7 days after booster vaccination (**Figure S3**). Most SARs were grade 1 or 2 with no grade 4 events reported. Pain (47.3%) was the most commonly reported local SAR; irritability/crying (53.2%), fatigue (33.3%), and loss of appetite (22.3%) were the most commonly reported systemic SARs. Fever (≥38⁰C/100.4⁰F) was reported by 41 participants (7.6%); grade 3 fever was reported by 5 participants (0.9%), all in the 2-to-5-year age group. Median (range) time of SAR onset was 1.0 (1.0-7.0) day and duration was 2.0 (1.0-14.0) days after booster.

Unsolicited AEs were reported by 22.3% of participants within 28 days after booster (**Table S5**); 14 (2.6%) were considered vaccine-related. The most reported unsolicited AE was upper respiratory tract infection (5.8%) and the most common unsolicited AEs considered vaccine-related were diarrhea, vomiting, dermatitis, and urticaria (0.4% each). One participant (0.2%) experienced an AESI considered vaccine-related 1 day after booster (erythema multiforme, mild); of note, topical silver sulfadiazine was used for the treatment of bacterial folliculitis previously by this participant. No SAEs, fatal events, or study discontinuations due to AEs occurred within 28 days following booster. Five SAEs and an additional AESI occurred up to the data cutoff, none of which were considered vaccine-related.

#### Immunogenicity

In the primary analysis (PPIS-Neg), booster vaccination induced nAb responses against BA.1 that were superior to those following mRNA-1273 primary series in KidCOVE (**Figure 3A**). The observed nAb GMCs (95% CI) against BA.1 were 822.0 (737.4-916.5) after mRNA-1273.214 booster and 65.7 (60.6-71.3) after dose 2 of mRNA-1273, respectively; the estimated GMR was 12.5 (95% CI, 11.0-14.3), meeting the pre-specified superiority criterion. Seroresponse rate against BA.1 relative to pre-dose 1 was 99.7% (95% CI, 98.1-100.0) after mRNA-1273.214 booster and 86.1% (95% CI, 81.1-90.2) after mRNA-1273 primary series (**Figure 3B**). The SRR difference was 13.6% (95% CI, 9.7-18.5), meeting the pre-specified non-inferiority criterion.

**Figure 3.**
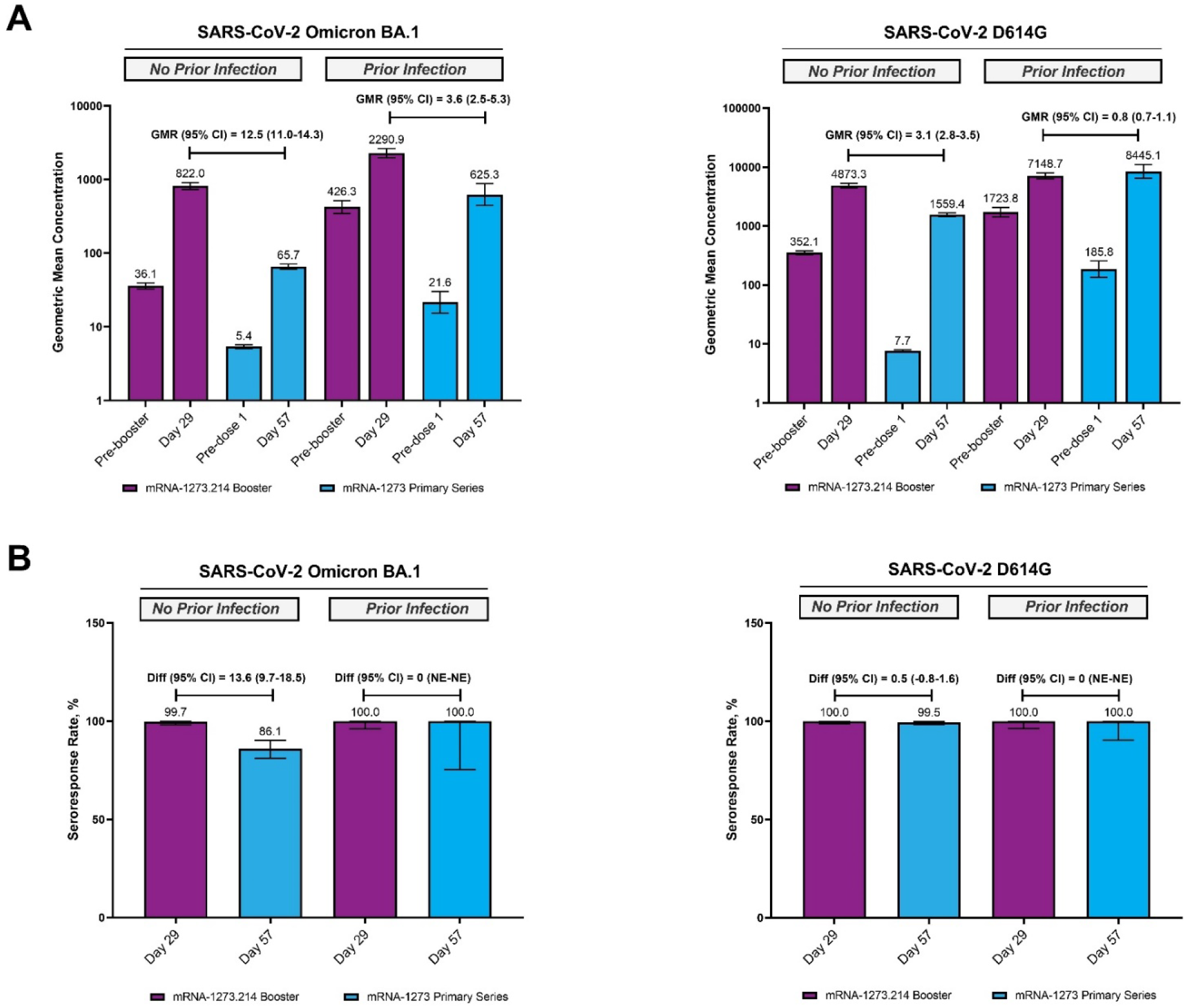
Serum neutralizing antibodies against BA.1 and D614G (A) and seroresponse rates (B) after mRNA-1273.214 booster or mRNA-1273 primary series vaccination in Part 2 Pseudovirus nAbs against BA.1 or D614G were measured before the mRNA-1273.214 booster dose (pre-booster) and at 28 days after the booster dose (Day 29) among participants in the PPIS, as well as before the mRNA-1273 primary series (pre-dose 1) and 28 days after the mRNA-1273 primary series dose 2 (Day 57). No prior SARS-CoV-2 infection was defined as negative RT-PCR test and negative Elecsys result on or before Day 1. Prior SARS-CoV-2 infection was indicated by a positive RT-PCR test or positive Elecsys result on or before Day 1. No prior infection: mRNA-1273.214 booster, n=299; mRNA-1273, n=590; prior infection: mRNA-1273.214 booster, n=136; mRNA-1273, n=42. Serologic response was defined as an increase in antibody levels from below the LLOQ at baseline to ≥4 times the LLOQ, or a ≥4-fold rise if baseline was equal or above the LLOQ. Antibody values below the LLOQ were replaced by 0.5 x LLOQ. Values greater than the ULOQ were replaced by the ULOQ if actual values were not available. LLOQs were 8 (BA.1) or 10 (D614G); ULOQs were 41984 (BA.1) or 4505600 (D614G). CI, confidence interval; GMC, geometric mean concentration; GMR, geometric mean ratio; LLOQ, lower limit of quantification; nAb, neutralizing antibody; PPIS, per-protocol immunogenicity set; RT-PCR, reverse transcription-polymerase chain reaction; ULOQ, upper limit of quantification.

Neutralizing antibody responses against D614G after booster vaccination were non-inferior to those induced by mRNA-1273 primary series in KidCOVE (**Figure 3A**). Among the PPIS-Neg, observed nAb GMCs (95% CI) against D614G were 4873.3 (4453.6-5332.5) and 1559.4 (1459.2-1666.6) after mRNA-1273.214 booster and mRNA-1273 primary series, respectively. The estimated GMR of 3.1 (95% CI, 2.8-3.5) met the pre-specified non-inferiority criterion. SRRs relative to pre-dose 1 were 100.0% (95% CI, 98.7-100.0) and 99.5% (95% CI, 98.4-99.9) for mRNA-1273.214 boosted participants and mRNA-1273 primary series recipients, respectively (**Figure 3B**). The SRR difference of 0.5% (95% CI, –0.8 to -1.6) met the pre-specified non-inferiority criterion.

## Discussion

The omicron-BA.1 variant-containing vaccine (mRNA-1273.214), administered as a 2-dose primary series (25 µg) or a booster (10 µg), had an acceptable safety profile with no new safety concerns in children aged 6 months to 5 years. Compared to mRNA-1273 primary series, elicited nAb responses were superior against BA.1 and non-inferior against D614G. Overall, these interim findings are consistent with current data on variant-containing vaccination in adults. This study is among the first to demonstrate the safety and immunogenicity of a variant-containing COVID-19 vaccine for use as a primary series and booster in pediatric populations, serving as a model of this approach in children.

The safety profile of mRNA-1273.214 as a primary series and booster dose was consistent with the mRNA-1273 primary series in this age group.^7^ Unsolicited AEs were generally reflective of common pediatric illnesses, and there were no vaccine-related SAEs or severe AEs. Reactogenicity profiles of the primary series and booster were characterized by mild-to-moderate, transient reactogenicity across age cohorts, with few grade 3, and no grade 4 reactions observed. Overall, no new safety concerns were identified, and both the primary series and booster had an acceptable reactogenicity profile in children aged 6 months to 5 years.

An omicron-BA.4/BA.5 variant-containing vaccine (mRNA-1273.222) is currently authorized in the United States as a 2-dose primary series in children aged 6 months to 5 years,^11, 17^ as this population, and younger children in particular, have lower SARS-CoV-2 seropositivity than older age groups.^18^ Development of variant-containing formulations suitable for primary vaccination in children is a necessary response to an anticipated reduction in seropositivity among children, with aging-in of new birth cohorts paired with continued SARS-CoV-2 evolution. In this study, the effectiveness of a mRNA-1273.214 primary series in children was inferred from meeting pre-specified success criteria for immunobridging to age-matched historical controls who received mRNA-1273 primary series in KidCOVE.^7^ Overall, these data suggest that variant-containing primary vaccination will potentially better protect young children as SARS-CoV-2 variants evolve and establishes a proof of principle for this approach among children.

The omicron-BA.4/BA.5 variant-containing vaccine is also authorized in the United States as a booster dose in children aged 6 months to 5 years.^11, 17^ In this study, evaluation of an omicron-BA.1 variant-containing vaccine (mRNA-1273.214) booster after completing the original primary series provided insights into variant responses in children. Effectiveness of the booster was inferred based on meeting pre-specified success criteria for immunobridging to mRNA-1273 primary series recipients in KidCOVE.^7^ Our findings in young children are consistent with those in adults, with omicron-containing booster vaccination eliciting superior immune responses against vaccine-matched BA.1 or BA.4/BA.5.^12, 13^ Moreover, a BA.4/BA.5-containing booster (mRNA-1273.222) in adults induced nAbs against BQ.1.1 and XBB.1 subvariants, highlighting the breadth afforded by variant-containing formulations.^14, 19^ Other studies of mRNA-based COVID-19 vaccines have shown persistence of germinal center B cell responses, enabling robust humoral immunity against SARS-CoV-2.^20, 21^ Additionally, COVID-19 incidence rates trended lower among mRNA-1273.214 booster recipients than mRNA-1273 booster recipients in a clinical trial of individuals aged ≥16 years.^13^ Notably, real-world studies showed variant-containing boosters provide additional protection against symptomatic infection^22^ or COVID-19–associated hospitalization^23^ among adults who received ≥2 doses of a vaccine targeting the ancestral strain. Overall, the observed benefits of booster vaccination in adults are expected to extend to young children. Further, given that efficacy of the original vaccine declined with the emergence of a new immune-evading variant,^6^ ongoing booster vaccinations would only be anticipated when vaccine formulation is updated to better match emergent SARS-CoV-2 variants.

Trial limitations include the open-label study design and use of a non-contemporaneous comparative cohort. The inclusion of a placebo-controlled or an active-controlled arm was precluded due to established COVID-19 vaccination recommendations in this age group. Additionally, this interim analysis presents data from a small population of Part 1 participants who had a higher percentage of baseline SARS-CoV-2–positive status than previously observed among KidCOVE participants, which could potentially impact immunogenicity comparisons. All participants were included for the primary analysis since the cohort is reflective of the serologic status of the unvaccinated population. When analyses were limited to baseline SARS-CoV-2– negative participants, substantially higher nAb concentrations were still observed against BA.1 after mRNA-1273.214 versus mRNA-1273 vaccination. However, nAb concentrations against ancestral SARS-CoV-2 among baseline SARS-CoV-2 negative participants were lower after mRNA-1273.214 versus mRNA-1273. But as SARS-CoV-2 has evolved, data demonstrate that a variant-containing vaccine more closely matched to circulating strains is more beneficial than the original vaccine.^13^ Further studies of variant-containing vaccines in children are ongoing. A phase 2 trial evaluating the safety, tolerability, and effectiveness of mRNA-1273.214 among infants aged 12 weeks to 5 months is recruiting (NCT05584202).

## Conclusion

In children aged 6 months to 5 years, a 2-dose 25-µg primary series and 10-µg booster dose of mRNA-1273.214 were immunogenic against BA.1 and D614G, with a comparable safety profile to mRNA-1273 primary vaccination. These findings are among the first to demonstrate the safety and inferred effectiveness of a variant-containing COVID-19 vaccine in pediatric populations and support a 2-dose schedule for previously unvaccinated children with an additional dose when the formulation is successively updated. Further studies are needed to assess the effectiveness and durability of variant-containing COVID-19 vaccines in children.

## Funding/Support

This work was supported by Moderna, Inc.

## Supporting information

Consort Checklist

## Data Availability

As the trial is ongoing, access to patient-level data presented in the article (immunogenicity, safety, and reactogenicity) and supporting clinical documents by qualified external researchers who provide methodologically sound scientific proposals may be available upon reasonable request and subject to review once the trial is complete. Such requests can be made to Moderna, Inc., 200 Technology Square, Cambridge, MA 02139. A materials transfer and/or data access 343 agreement with the sponsor will be required for accessing shared data. All other relevant data are presented in the paper.

## Acknowledgments

We thank the participants and their families for their involvement in the trial, as well as the staff at the trial sites for implementing the protocol. Medical writing and editorial assistance were provided by Jared Mackenzie, PhD, Kate Russin, PhD, and Lindsey Kirkland, PhD, of MEDiSTRAVA in accordance with Good Publication Practice guidelines, funded by Moderna, Inc., and under the direction of the authors.

## Conflicts of Interest Disclosures (includes financial disclosures)

AD, RP, KH, BG, WZ, WD, HZ, SSG, JMM, and RD are employees of Moderna, Inc., and hold stock/stock options in the company. AY, KS and BJK are consultants for Moderna, Inc.

## Data Sharing Statement

As the trial is ongoing, access to patient-level data presented in the article (immunogenicity, safety, and reactogenicity) and supporting clinical documents by qualified external researchers who provide methodologically sound scientific proposals may be available upon reasonable request and subject to review once the trial is complete. Such requests can be made to Moderna, Inc., 200 Technology Square, Cambridge, MA 02139. A materials transfer and/or data access agreement with the sponsor will be required for accessing shared data. All other relevant data are presented in the paper.

## Author Contributions

A.D., R.B., K.A., M.T., K.H., K.S., W.D., H.Z., S.S.G., J.M.M., F.P., and R.D. conceived and designed the current study. A.D., R.B., K.A., C.G., R.A.C., M.T., R.P., B.K., K.S., A.Y., B.G., W.Z., W.D., H.Z., S.S.G., J.M.M., F.P., and R.D. carried out analysis and interpretation of data and take responsibility for the integrity of the data and accuracy of the data analysis. All authors critically reviewed the paper for important intellectual content and approved the final draft of the manuscript.

## Notes

### Clinical Trial

NCT05436834

### Author Declarations

The protocol and study documents were approved before conduct of study procedures (Advarra Institutional Review Board). Written informed consent from parent(s)/legally authorized representative(s) of children was obtained before performing study procedures. The study was conducted in accordance with the protocol, ethical principles derived from international guidelines that including the Declaration of Helsinki and Council for International Organizations of Medical Sciences International Ethical Guidelines, International Council for Harmonisation Good Clinical Practice guidelines, and laws and regulatory requirements.

