## Supplementary Information for "Interim Safety and Immunogenicity of COVID-19 Omicron-BA.1 Variant-Containing Vaccine in Children"

**Supplement**

***List of ROVER Study Investigators (Enrolled Before December 5, 2022)***

| Site Number | Principal Investigator | Location Name | Address |
|---|---|---|---|
| US002 | Ali, Kashif | Texas Center for Drug Development | 6550 Mapleridge St, Ste 201, Houston, Texas, United States, 77081 |
| US007 | Ampajwala, Madhavi | Village Health Partners Frisco Medical Village | 9990 Dallas Parkway, Suite 200, Frisco, Texas, United States, 75033 |
| US030 | Atz, Andrew | MUSC | 169 Ashley Avenue, MH Rm 161, Charleston, South Carolina, United States, 29425 |
| US045 | Bennett, Richard | Clinical Research Partners LLC | 7110 Forest Ave Ste 201, Richmond, Virginia, United States, 23226 |
| US003 | Berman, Gary | Clinical Research Institute, Inc - CRN - PPDS | 825 Nicollet Mall Ste 1135, Minneapolis, Minnesota, United States, 55402-2700 |
| US025 | Clifford, Robert | Coastal Pediatric Associates | 2051 Charlie Hall Blvd, Charleston, South Carolina, United States, 29414-5834 |
| US035 | Dunn, Michael | Quality Clinical Research - HyperCore - PPDS | 10040 Regency Cir, Suite 375, Omaha, Nebraska, United States, 68114 |
| US088 | Eder, Frank | Meridian Clinical Research (Binghamton-New York) - PPDS | 1290 Upper Front St, Binghamton, New York, United States, 13901-1046 |
| US071 | Fuchs, George | University of Kentucky | 1000 South Limestone, Room A03.101, Lexington, Kentucky, United States, 40536 |
| US083 | Griffin, Carl | Lynn Health Science Institute - ERN - PPDS | 3555 North West 58th Str Suite 800, Oklahoma City, Oklahoma, United States, 73112 |
| US032 | Hernandez, John M. | Sera Collection Research Services, LLC | 3317 W. Beverly Blvd., Montebello, California, United States, 90640 |
| US064 | Jeanfreau, Robert | MedPharmics, LLC | 3800 Houma Boulevard, Suite 335A, Metairie, Louisiana, United States, 70006 |
| US041 | Johnson, Kimball | IResearch Atlanta LLC | 250 E Ponce De Leon Ave Ste 800, Decatur, Georgia, United States, 30030-3438 |

| | | | |
|---|---|---|---|
| US094 | Lavery, William | Meridian Clinical Research (Overland Park, Kansas) - PPDS | 8675 College Boulevard, Suite 200, Overland Park, Kansas, United States, 66210 |
| US106 | Meyer, Jay | Meridian Clinical Research, LLC | 4600 Valley Road, Lincoln, Nebraska, United States, 68510 |
| US033 | Muller, William | Ann and Robert H. Lurie Childrens Hospital of Chicago | 225 East Chicago Avenue Room 19-371, Attn: IDS Pharmacy, Chicago, Illinois, United States, 60611-2605 |
| US016 | Ohnmacht, Richard | Velocity Clinical Research-Providence | 1598 South County Trail, Suite 204, East Greenwich, Rhode Island, United States, 02818 |
| US049 | Padhye, Amruta | University of Missouri Health Care System | 1 Hospital Drive Room T0024B, Columbia, Missouri, United States, 65212 |
| US004 | Palanpurwala, Khozema | Cyfair Clinical Research Center - ERN - PPDS | 11830 FM 1960 W, Houston, Texas, United States, 77065 |
| US067 | Rodriguez, Carina | University of South Florida | USF CIRP, 13330 USF Laurel Drive, Room 6116, Tampa, Florida, United States, 33612 |
| US021 | Turner, Mark | Velocity Clinical Research - Boise - ERN - PPDS | 2950 E Magic View Dr Ste 182, Meridian, Idaho, United States, 83642 |
| US097 | Vasko, Todd | Meridian Clinical Research | 3030 Ashley Town Center Dr, Suite 102A, Charleston, South Carolina, United States, 29414 |
| US096 | Waits, John | Trinity Clinical Research, LLC | 975 9th Ave. S.W., Ste 300, Bessemer, Alabama, United States, 35022 |
| US095 | Warfield, Peter | Meridian Clinical Research (Washington) - PPDS | 4910 Massachusetts Ave NW, Suite 315, Washington, District of Columbia, United States, 20016 |

***Inclusion/Exclusion Criteria***

Participants, including those who are currently enrolled in KidCOVE, are eligible to be included in this study only if all the following criteria apply:

1. The participant is male or female, aged 6 months to 5 years, at the time of consent (Screening Visit), who is in good general health, in the opinion of the investigator, based on review of medical history and screening physical examination. Note: Participant must be aged ≥6 months and must not have completed 6 years of age at the time of administration of first dose.
2. If the participant has a chronic disease (eg, asthma, diabetes mellitus, cystic fibrosis, HIV infection), the disease should be stable, per investigator assessment, so that the participant can be considered eligible for inclusion. Stable diseases are those which have had no change in their status or in the medications required to control them in the 6 months prior to Screening Visit.

   Note: A change in medication for dose optimization (eg, insulin dose changes, adjustments for age-related weight gain), change within class of medication, or reduction in dose are not considered signs of instability.
3. In the investigator's opinion, the parent(s)/LAR(s) understand and are willing and physically able to comply with protocol-mandated follow-up, including all procedures and provide written informed consent. This includes inability to draw baseline blood samples (minimum amount needed).
4. The participant is aged ≥2 years and has a BMI at or above the 2nd percentile according to WHO Child Growth Standards at the Screening Visit.

   OR

The participant is aged <2 years and the participant's height and weight are both at or above the 2nd percentile according to WHO Child Growth Standards at the Screening Visit.

Special inclusion criterion for participants aged 6 to <12 months:

5. The participant was born at full-term (≥37 weeks gestation) with a minimum birth weight of 2.5 kg.

Inclusion criterion for Part 2:

6. The participant must have received 2 doses of mRNA-1273, approximately 28 to 35 days apart, as 25-µg primary series, and second dose was given ≥4 months prior to enrollment.

Participants will be excluded from the study if any of the following criteria apply:

1. Has a known history of SARS-CoV-2 infection (eg, reported AE of COVID-19 or asymptomatic SARS-CoV-2 infection during KidCOVE) in the 90 days prior to dosing in this study.
2. Is acutely ill or febrile 24 hours prior to or at the Screening Visit. Fever is defined as a body temperature ≥38.0°C/≥100.4°F. Participants who meet this criterion may have visits rescheduled within the relevant study visit windows. Afebrile participants with minor illnesses can be enrolled at the discretion of the investigator.
3. Has previously been administered an investigational or approved CoV (eg, SARS-CoV-2, SARS-CoV, MERS-CoV) vaccine. For Part 2, this applies to vaccines other than the mRNA-1273 (prototype) vaccine.

4. Has undergone treatment with investigational or approved agents for prophylaxis against COVID-19 (including receipt of SARS-CoV-2 monoclonal antibodies for prophylaxis or treatment) within 90 days prior to enrollment.

5. Has a known hypersensitivity to a component of the vaccine or its excipients. Hypersensitivity includes, but is not limited to, anaphylaxis or immediate allergic reaction of any severity to a previous dose of an mRNA COVID-19 vaccine or any of its components (including PEG or immediate allergic reaction of any severity to polysorbate).

6. Has a medical or psychiatric condition that, according to the investigator's judgment, may pose additional risk as a result of participation, interfere with safety assessments, or interfere with interpretation of results.

7. Has a history of diagnosis or condition that, in the judgment of the investigator, may affect study endpoint assessment or compromise participant safety, specifically the following:

- Congenital or acquired immunodeficiency, other than well-controlled HIV infection as described in Inclusion Criterion 2
- Chronic hepatitis or suspected active hepatitis
- A bleeding disorder that is considered a contraindication to intramuscular injection or phlebotomy
- Dermatologic conditions that could affect local SAR assessments
- Any prior diagnosis of malignancy (excluding nonmelanoma skin cancer)

8. Has received the following:

• Any routine vaccination with inactivated or live vaccine(s) within 14 days prior to study vaccination or plans to receive such a vaccine through 14 days following study vaccination.

Note: This excludes influenza vaccine that may be given any time, but ideally ≥7 days before study dose. If a participant receives an influenza vaccine, this should be captured within the concomitant medication electronic case report form (eCRF).

• Systemic immunosuppressants or immune-modifying drugs for >14 days in total within 6 months prior to the day of enrollment (for corticosteroids, ≥1 mg/kg/day or ≥10 mg/day prednisone equivalent, if participant weighs >10 kg). Participants may have visits rescheduled for enrollment if they no longer meet this criterion within the Screening Visit window. Inhaled, nasal, and topical steroids, and palivizumab are allowed.

• Intravenous or subcutaneous blood products (red cells, platelets, immunoglobulins) within 3 months prior to enrollment.

9. Has participated in an interventional clinical study within 28 days prior to the Screening Visit or plans to do so while participating in this study (Note: The exception would be participants who rollover from KidCOVE into this study).

10. Is an immediate family member, or household contact, of an employee of the study site or Moderna or someone otherwise directly involved with the conduct of the study. As applicable, family members/household contacts of employees of the larger institution or affiliated private practice not part of the study site may be enrolled, as long as the

investigator has no authority over the family member's employment or performance evaluation.

11. Is currently experiencing a SAE in KidCOVE at the time of screening for this study.

#### *Ethics Statement*

The study was conducted in accordance with the protocol, ethical principles derived from international guidelines that including the Declaration of Helsinki and Council for International Organizations of Medical Sciences International Ethical Guidelines, International Council for Harmonisation Good Clinical Practice guidelines, and laws and regulatory requirements.

#### *Objectives*

Secondary objectives for Part 1 included evaluating immune responses against BA.1 and D614G induced by the mRNA-1273.214 primary series versus the mRNA-1273 primary series.

#### *Sample Size Calculations*

Parts 1 and 2 sample sizes were considered sufficient to support the safety evaluation of the mRNA-1273.214 vaccine in participants aged 6 months to 5 years, with approximately 480 participants each enrolled to receive the mRNA-1273.214 primary series in Part 1 and the mRNA-1273.214 booster in Part 2. The study had ≥90% probability to observe ≥1 participant with an AE at a true 0.5% AE rate in Parts 1 and 2. The sample sizes required to evaluate effectiveness of the mRNA-1273.214 primary series and booster by immunobridging to the mRNA-1273 primary series in the same age group in KidCOVE was calculated based on the superiority hypothesis testing of the serum nAb GMC for Parts 1 and 2. Additionally, the calculated sample size had adequate power (>90%) to perform noninferiority testing for the SRR.

For superiority hypothesis testing in Part 1, nAb GMCs against BA.1 after the mRNA-1273.214 primary series compared with nAb GMCs against BA.1 after the mRNA-1273 primary series in KidCOVE, assuming the true antibody GMR of 1.5 at Day 57, with a superiority margin of 1.0 and the standard deviation (SD) of the natural log-transformed levels of 1.8, 416 participants in PPIS were needed to provide 90% power at 1-sided α of 0.025. With the consideration of some participants excluded from the PPIS, the Part 1 target enrollment was approximately 480 participants.

For superiority hypothesis testing in Part 2, nAb GMCs against BA.1 after the mRNA-1273.214 booster dose compared with nAb GMCs against BA.1 after the mRNA-1273 primary series in KidCOVE, assuming the true antibody GMR of 1.5 (GMC at booster dose Day 29 after the mRNA-1273.214 booster compared with GMC at Day 57 after mRNA-1273 primary series in KidCOVE), with a superiority margin of 1.0 and the SD of the natural log-transformed levels of 1.5, 289 participants in PPIS-Neg were needed to provide 90% power at 1-sided α of 0.025.

The calculated sample size of 289 participants in the PPIS-Neg provided adequate power to perform the noninferiority hypothesis testing for the SRR against BA.1 after the mRNA-1273.214 booster dose compared with SRR against BA.1 after the mRNA-1273 primary series in KidCOVE. Assuming 80% SRR after the mRNA-1273 primary series in KidCOVE, and the true SRR difference of 10%, with a margin of -5%, the testing power will be >90% to claim noninferiority of SRR at 1-sided α of 0.025. With the consideration of some participants excluded from the PPIS-Neg, the target enrollment of Part 2 was approximately 480 participants.

***Immunogenicity Analyses***

SRR differences with 95% CIs (Miettinen-Nurminen score method) were determined for BA.1 and D614G in a primary analysis after booster in Part 2 and a secondary analysis after primary series in Part 1. Noninferiority was declared for BA.1 if the LB of the SRR difference 95% CI was >–5% and was declared for D614G if the LB of the difference 95% CI was >–10%.

### Supplemental Figures

Figure S1. Overview of the ROVER study design

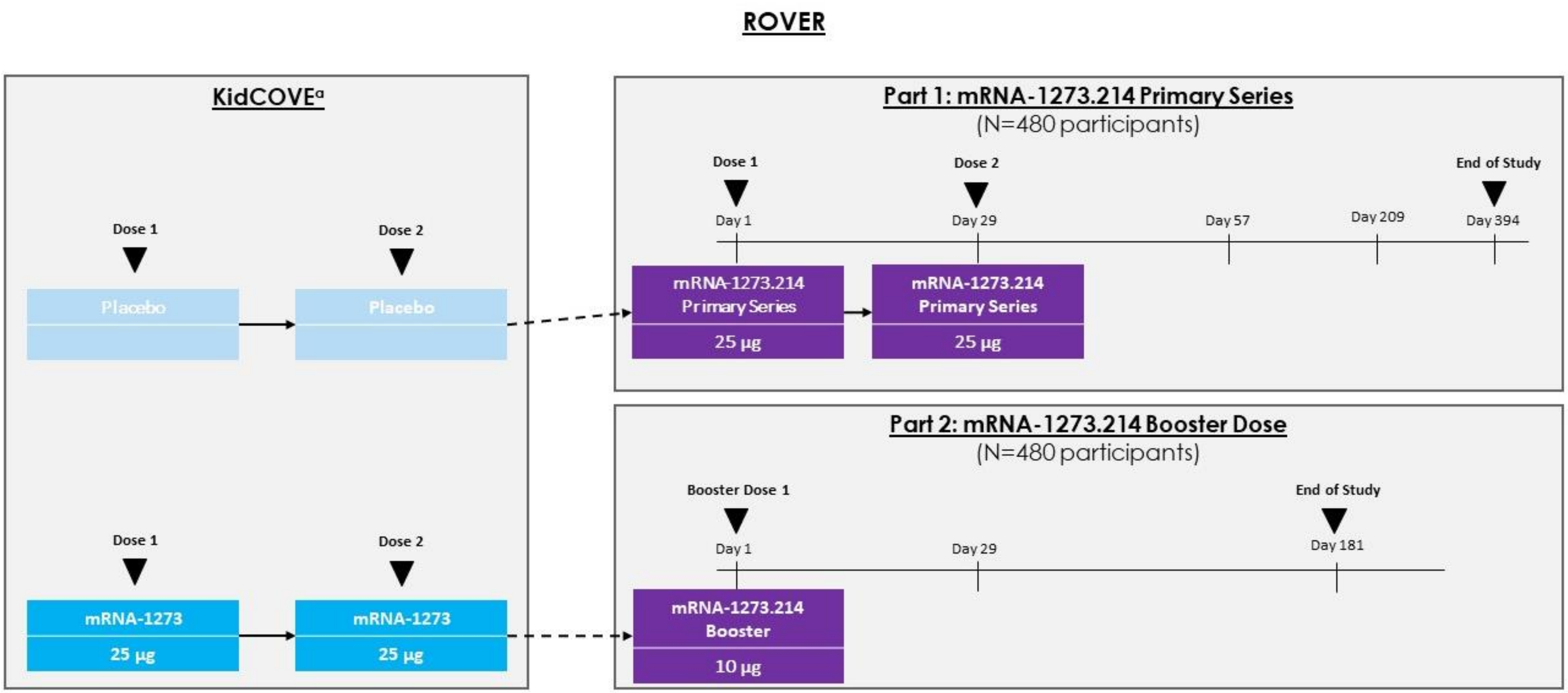


[a]Participants in the KidCOVE trial were eligible for inclusion in Part 1 or Part 2 of the ROVER trial. Part 1 enrolled participants who were not previously vaccinated against SARS-CoV-2, including placebo recipients from KidCOVE; Part 2 enrolled mRNA-1273 recipients from KidCOVE.

**Figure S2.** (A) Local solicited adverse reactions among participants aged 6 months to 5 years[a] and (B) systemic solicited adverse reactions among participants aged 6-36 months and 37 months to 5 years[b] within 7 days after each dose of the mRNA-1273.214 primary series in Part 1 (solicited safety set)

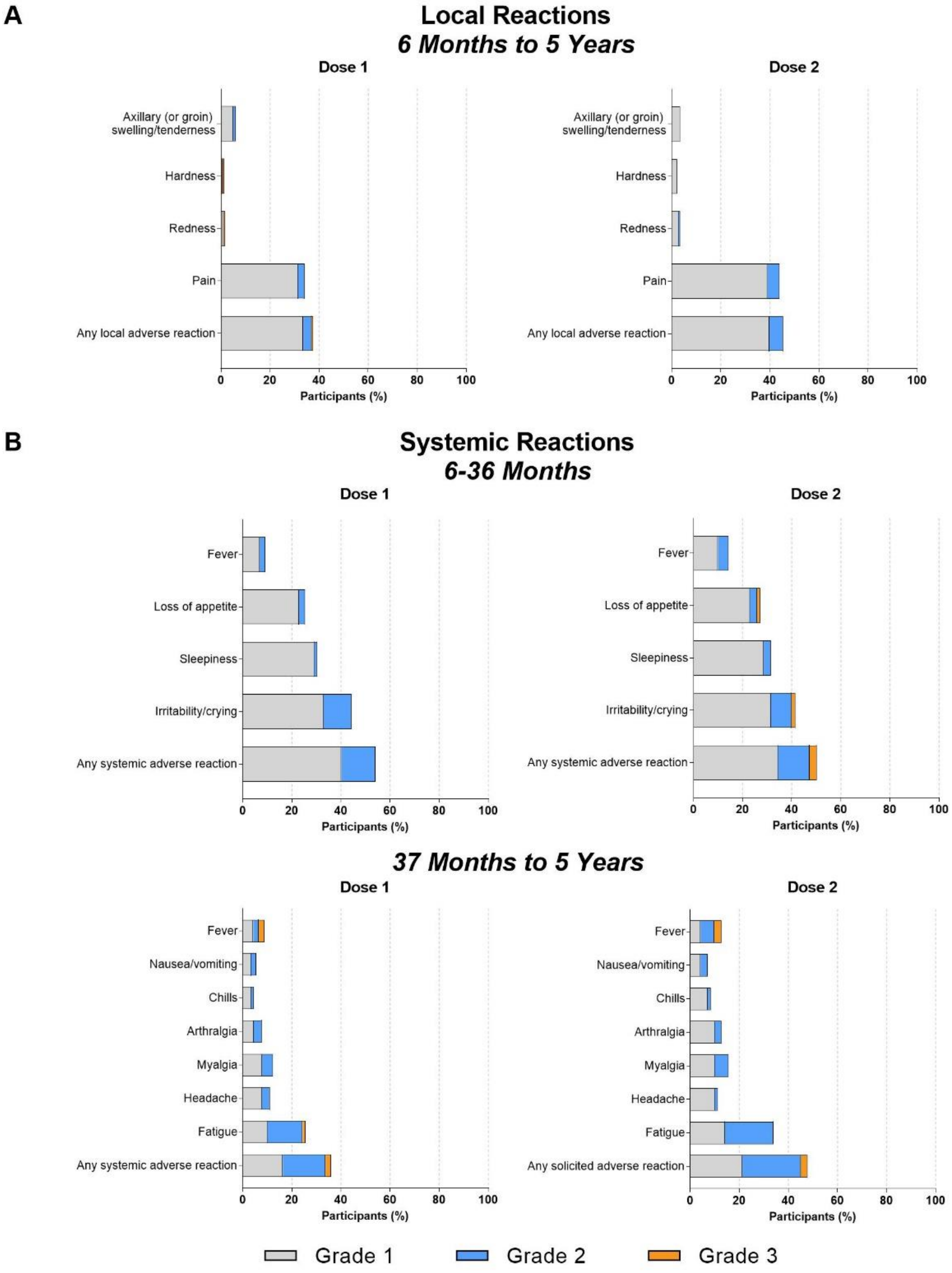

[a]Local adverse reactions collected among participants aged 6 months to 5 years were injection site pain, redness (erythema), hardness, and axillary (or groin) swelling/tenderness.

[b]Systemic adverse reactions collected among participants aged 6-36 months were fever, irritability/crying, sleepiness, and loss of appetite. Systemic adverse reactions collected among participants aged 37 months to 5 years were headache, fatigue, myalgia, arthralgia, nausea/vomiting, chills, and fever.

Percentages based on the number of exposed participants who submitted any data for the event.

Local adverse reactions: 6 months to 5 years, dose 1, n=179; 6 months to 5 years, dose 2, n=141.

Systemic adverse reactions: 6-36 months, dose 1, n=87; 37 months to 5 years, dose 1, n=92; 6-36 months, dose 2, n=70; 37 months to 5 years, dose 2, n=71.

**Figure S3.** (A) Local solicited adverse reactions among participants aged 6 months to 5 years[a] and (B) systemic solicited adverse reactions among participants aged 6-36 months and 37 months to 5 years[b] within 7 days after the mRNA-1273.214 booster dose in part 2 (solicited safety set)

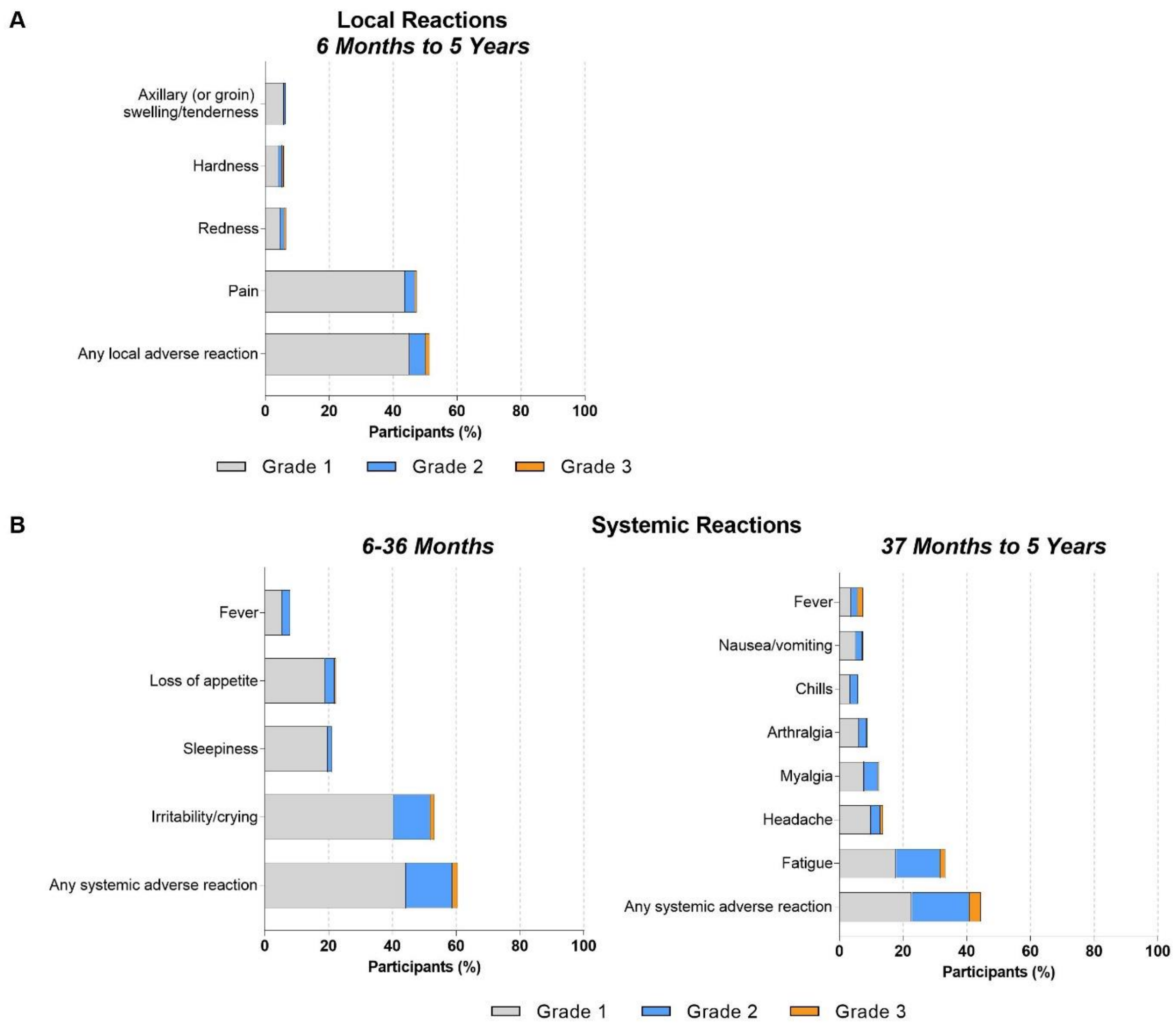


[a]Local adverse reactions collected among participants aged 6 months to 5 years were injection site pain, redness (erythema), hardness, and axillary (or groin) swelling/tenderness.

[b]Systemic adverse reactions collected among participants aged 6-36 months were fever, irritability/crying, sleepiness, and loss of appetite. Systemic adverse reactions collected among

participants aged 37 months to 5 years were headache, fatigue, myalgia, arthralgia, nausea/vomiting, chills, and fever.

Percentages based on the number of exposed participants who submitted any data for the event.

Local adverse reactions: 6 months to 5 years, n=539. Systemic adverse reactions: 6-36 months, n=240; 37 months to 5 years, n=299.

### Supplemental Tables

**Table S1.** Adverse Events of Special Interest

| Medical Concept | Medical Concept Descriptions/Guidance |
|---|---|
| **Anosmia, ageusia** | New onset of anosmia or ageusia associated with COVID-19 or idiopathic etiology<br>DOES NOT INCLUDE anosmia or ageusia associated with sinus/nasal congestion, congenital, or traumatic etiologies |
| **Subacute thyroiditis** | Acute inflammatory disease of the thyroid (immune-mediated or idiopathic)<br>DOES NOT INCLUDE new onset of chronic thyroiditis |
| **Acute pancreatitis** | New onset of pancreatitis in the absence of a clear, alternate etiology, such as alcohol, gallstones, trauma, recent invasive procedure, etc. |
| **Appendicitis** | Any event of appendicitis |
| **Rhabdomyolysis** | New onset of rhabdomyolysis in the absence of a clear, alternate etiology, such as drug/alcohol abuse, excessive exercise, trauma, etc. |
| **Acute respiratory distress syndrome** | New onset of acute respiratory distress syndrome/respiratory failure due to acute inflammatory lung injury<br>DOES NOT INCLUDE nonspecific symptoms of shortness of breath or dyspnea, nor events with underlying etiologies of heart failure or fluid overload |
| **Coagulation disorders** | New onset of thrombosis, thromboembolic event, or nontraumatic hemorrhage/bleeding disorder (ex. stroke, deep vein thrombosis, pulmonary embolism, disseminated intravascular coagulation, etc.) |
| **Acute cardiovascular injury** | New onset of clinically confirmed, acute cardiovascular injury, such as myocarditis, pericarditis, arrhythmia confirmed by electrocardiogram (ex. atrial fibrillation, atrial flutter, supraventricular tachycardia), stress cardiomyopathy, heart failure, acute coronary syndrome, myocardial infarction, etc.<br>DOES NOT INCLUDE transient sinus tachycardia/bradycardia, nonspecific symptoms such as palpitations, racing heart, heart fluttering or pounding, irregular heartbeats, shortness of breath, chest pain/discomfort, etc. |

| | |
|---|---|
| **Acute kidney injury** | New onset of acute kidney injury or acute renal failure in the absence of a clear, alternate etiology, such as urinary tract infection/urosepsis, trauma, tumor, nephrotoxic medications/substances, etc.<br>Increase in serum creatinine by ≥0.3 mg/dL (or ≥26.5 μmol/L) within 48 hours; OR increase in serum creatinine to ≥1.5 times baseline, known or presumed to have occurred within prior 7 days |
| **Acute liver injury** | New onset in the absence of a clear, alternate etiology, such as trauma, tumor, hepatotoxic medications/substances, etc.: > 3-fold elevation above the upper normal limit for alanine aminotransferase or aspartate aminotransferase; OR > 2-fold elevation above the upper normal limit for total serum bilirubin or gamma glutamyl transferase or alkaline phosphatase |
| **Dermatologic findings** | Chilblain-like lesions<br>Single organ cutaneous vasculitis<br>Erythema multiforme<br>Bullous rash<br>Severe cutaneous adverse reactions, such as Stevens-Johnson syndrome, Toxic epidermal necrolysis, drug reaction with eosinophilia and systemic symptoms, fixed drug eruptions, and necrotic or exfoliative reactions |
| **Systemic inflammatory syndromes** | Multisystem inflammatory syndrome in adults or multisystem inflammatory syndrome in children<br>Kawasaki's disease<br>Hemophagocytic lymphohistiocytosis |
| **Thrombocytopenia** | Platelet count <150 x $10^9$/L (thrombocytopenia)<br>New clinical diagnosis, or worsening, of thrombocytopenic condition, such as immune thrombocytopenia, thrombocytopenic purpura, or<br>HELLP syndrome (hemolysis, elevated liver enzymes, and low platelet count) |
| **Acute aseptic arthritis** | Clinical syndrome characterized by acute onset of signs and symptoms of joint inflammation without recent trauma for a period of no longer than 6 weeks, synovial increased leukocyte count and the absence of microorganisms on Gram stain, routine culture and/or polymerase chain reaction<br>DOES NOT INCLUDE new onset of chronic arthritic conditions |

| | |
|---|---|
| **New onset, or worsening, of neurological disease** | Immune-mediated neurological disorders<br>Guillain-Barre syndrome<br>Acute disseminated encephalomyelitis<br>Peripheral facial nerve palsy (Bell’s palsy)<br>Transverse myelitis<br>Encephalitis/encephalomyelitis<br>Aseptic meningitis<br>Seizures/convulsions/epilepsy<br>Narcolepsy/hypersomnia |
| **Anaphylaxis** | Anaphylaxis associated with study drug administration |
| **Other syndromes** | Fibromyalgia<br>Postural orthostatic tachycardia syndrome<br>Chronic fatigue syndrome<br>Myalgic encephalomyelitis<br>Post viral fatigue syndrome<br>Myasthenia gravis |

**Table S2.** Analysis Populations

| Analysis Set | Description |
|---|---|
| Full Analysis Set | Enrolled participants who receive ≥1 injection of investigational product. |
| Immunogenicity Set | Participants in the Full Analysis Set who provide immunogenicity samples. |
| PPIS | Participants in the Immunogenicity Set who receive planned doses of investigational product per schedule, comply with immunogenicity testing schedule, and have no major protocol deviations that impact key or critical data, regardless of prior SARS-CoV-2 infection status at the time of vaccination (primary series). Participants with HIV who are receiving HAART will be excluded. |
| PPIS-Neg | Participants in the PPIS who are SARS-CoV-2–negative (no serologic or virologic evidence of prior SARS-CoV-2 infection) before receiving the mRNA-1273.214 vaccine (pre-dose 1 for Part 1 or pre-booster dose for Part 2). |
| Safety Set | Enrolled participants who receive ≥1 dose of investigational product. |
| Solicited Safety Set | Participants in the Safety Set who contribute any SAR data. |

HAART, highly active antiretroviral therapy; PPIS, per-protocol immunogenicity set; PPIS-Neg, per-protocol immunogenicity set-negative; SAR, solicited adverse reaction.

**Table S3.** Summary of Unsolicited Treatment-Emergent Adverse Events Within 28 Days After Any Dose of the mRNA-1273.214 Primary Series in Part 1 (Safety Set)

| | **Total Participants (N=179)** |
|---|---|
| **Any TEAEs, n (%)** | |
| All | 55 (30.7) |
| SAE | 1 (0.6) |
| Fatal | 0 |
| MAAE | 45 (25.1) |
| Leading to discontinuation from study vaccine | 0 |
| Leading to discontinuation from study participation | 0 |
| Grade 3/severe | 1 (0.6) |
| Grade 3 or higher | 1 (0.6) |
| AESI | 0 |
| **Related TEAEs, n (%)** | |
| All | 2 (1.1)[a] |
| SAE | 0 |
| Fatal | 0 |
| MAAE | 0 |
| Leading to discontinuation from study vaccine | 0 |
| Leading to discontinuation from study participation | 0 |
| Grade 3/severe | 0 |
| Grade 3 or higher | 0 |
| AESI | 0 |

AE, adverse event; AESI, AE of special interest; MAAE, medically attended AE; SAE, serious AE; TEAE, treatment-emergent AE.

[a]The unsolicited TEAEs that were considered vaccine-related included diarrhea and croup.

**Table S4.** BA.1 and D614G Seroresponse Rates After the mRNA-1273.214 Primary Series Vaccination in Part 1 (Per-Protocol Immunogenicity Set)

| | **BA.1** | | **D614G** | |
|---|---|---|---|---|
| | **mRNA-1273.214 Primary Series** | **mRNA-1273 Primary Series** | **mRNA-1273.214 Primary Series** | **mRNA-1273 Primary Series** |
| | **n=71** | **n=632** | **n=71** | **n=632** |
| Day 57 SRR,[a] n/N (%) [95% CI] | 50/56 (89.3) [78.1-96.0] | 223/257 (86.8) [82.0-90.7] | 64/66 (97.0) [89.5-99.6] | 582/585 (99.5) [98.5-99.9] |
| Difference (95% CI)[b] | -1.6 (-11.3 to 8.2) | | -5.4 (-10.5 to -0.3) | |

CI, confidence interval; LLOQ, lower limit of quantification; SRR, seroresponse rate.

[a]Seroresponse at the participant level was defined as a change from baseline (pre-dose 1 of primary series) below the LLOQ to equal or above 4 x LLOQ; or ≥ a 4-fold rise if baseline was equal to or above the LLOQ and below 4 x LLOQ; or ≥ a 2-fold rise if baseline was equal to or above 4 x LLOQ. Percentages were based on N1. N1 = Number of participants with non-missing data at baseline and the corresponding timepoint. 95% CI was calculated using the Clopper-Pearson method.

[b]Common risk difference and 95% CI were calculated using the stratified Miettinen-Nurminen method adjusted by age group (6-23 months and 2-5 years).

**Table S5.** Summary of Unsolicited Treatment-Emergent Adverse Events Within 28 Days After the mRNA-1273.214 Booster Dose in Part 2 (Safety Set)

| | **Total Participants (N=539)** |
|---|---|
| **Any TEAEs, n (%)** | |
| All | 120 (22.3) |
| SAE | 0 |
| Fatal | 0 |
| MAAE | 79 (14.7) |
| Leading to discontinuation from study vaccine | 0 |
| Leading to discontinuation from study participation | 0 |
| Grade 3/severe | 0 |
| Grade 3 or higher | 0 |
| AESI | 1 (0.2) |
| **Related TEAEs, n (%)** | |
| All | 14 (2.6) |
| SAE | 0 |
| Fatal | 0 |
| MAAE | 3 (0.6) |
| Leading to discontinuation from study vaccine | 0 |
| Leading to discontinuation from study participation | 0 |
| Grade 3/severe | 0 |
| Grade 3 or higher | 0 |
| AESI | 1 (0.2) |

AE, adverse event; AESI, AE of special interest; MAAE, medically attended AE; SAE, serious AE; TEAE, treatment-emergent AE.
